# Unsupervised subgrouping of chronic low back pain patients treated in a specialty clinic

**DOI:** 10.1101/2023.11.04.23298104

**Authors:** Abel Torres-Espin, Anastasia Keller, Susan Ewing, Andrew Bishara, Naoki Takegami, Adam R. Ferguson, Aaron Scheffler, Trisha Hue, Jeff Lotz, Thomas Peterson, Patricia Zheng, Conor O’Neill

**Affiliations:** School of Public Health Sciences, University of Waterloo, ON, Canada; Department of Physical Therapy, University of Alberta, AB, Canada; Department of Neurosurgery, Brain and Spinal Injury Center, Weill Institutes for Neurosciences, University of California, San Francisco, CA, United States; Department of Epidemiology and Biostatistics, University of California, San Francisco, CA, United States; Department of Anesthesia and Perioperative Care, University of California, San Francisco, CA, United States; Bakar Computational Health Sciences Institute, University of California, San Francisco, CA, United States; San Francisco Veterans Affairs Healthcare System, San Francisco, CA, United States; Department of Orthopaedic Surgery, University of California, San Francisco, CA, United States

**Author notes:** co-corresponding authors Abel Torres Espin Patricia Zheng Conor O’Neill.

**Keywords:** Unsupervised learning, clustering, phenotyping, chronic low back pain, specialty clinic

## Abstract

**Background:** Chronic low back pain (cLBP) is the leading cause of disability worldwide. Current treatments have minor or moderate effects, partly because of the idiopathic nature of most cLBP cases, the complexity of its presentation, and heterogeneity in the population. Explaining this complexity and heterogeneity by identifying subgroups of patients is critical for personalized health. Clinical decisions tailoring treatment to patients’ subgroup characteristics and specific treatment responses can improve health outcomes. Current patient stratification tools divide cases into subgroups based on a small subset of characteristics, which may not capture many factors determining patient phenotypes.

**Methods and Findings:** In this study, we use an unsupervised machine learning framework to identify patient subgroups within a specialized back pain clinic and evaluate their outcomes. Our analysis identified 25 latent factors determining patient phenotypes and found three distinctive clusters of patients. The research suggests that there is heterogeneity in the population of patients treated in a specialty setting and that several factors determine patient phenotypes. Cluster 1 consists of those individuals with characteristics found to be protective of chronic pain: younger age, low pain medication prescription, high function, good insurance access, and low overlapping pain conditions. Individuals in Cluster 3 associate with older age and present with a higher incidence of chronic overlapping pain conditions, comorbidities, and pain medication use. Cluster 2 is an intermediate group.

**Conclusions:** We quantify cLBP population heterogeneity and demonstrate how ML analytical workflow can be used to explain, in part, this heterogeneity in relation to outcomes. Notably, considering a data-driven approach from multi-domain data produces different subgroups than the STarT back screening tool, and the addition of other functional metrics at baseline such as global physical and mental function, and pain intensity, increases the variance explained in outcomes. Our study provides novel insights into the complex nature of cLBP and the potential for data-driven methods to identify clinically relevant subtypes.

## Introduction

Chronic low back pain (cLBP) is the leading cause of disability worldwide (1). With a lifetime prevalence of 70%, cLBP affects over half of all Americans annually (1–3), yet effective non-opioid treatment remains elusive. cLBP is a complex pathology with a heterogeneous population (4,5), and several interrelated factors influencing its presentation and progression. About 90% of cases are considered non-specific, which means there are no clear causes for the presence of pain. As a consequence, heterogeneous responses to treatments are likely. One commonly used framework for understanding cLBP complexity is the biopsychosocial (BPS) model (6), in which cLBP results from an interplay between biological factors such as complex pain signal processing in the central nervous system (CNS), and psychological and social factors (7,8). For example, age, gender, race, culture, and comorbidities have been shown to associate with cLBP outcomes (9–14). While widely accepted, the results from treatments inspired by the BPS model of cLBP are typically small, with randomized clinical trials (RCT) demonstrating that even the best treatments improve pain by only two points on the 0-11 Visual Analog Scale (VAS) (7,15,16). RCT’s however, generally estimate an average treatment effect, which may obscure differences in treatment effects in subgroups of the population. Identifying these subgroups is critical for personalized health, where clinical decision tailoring treatment depending on a patient’s subgroup characteristics and specific response to treatment can improve health outcomes (17–19).

Several classification, stratification, and subgrouping tools have been developed to guide screening and clinical decision making (17–19). The evidence for their utility is often inconsistent. For example, studies using the Subgroup for Targeted Treatment Back Screening Tool (STarT back, or SBT), for stratified care management has shown contradictory results, with studies describing positive outcomes (20,21) while others reporting inconclusive effects (22,23). A remaining question is whether SBT is sufficient or whether other stratifying factors can add valuable information on subgrouping patients for risk prediction and treatment response (24). These subgrouping strategies are often derived from a small subset of metrics capturing a focused aspect of pain experience, and based largely on expert opinion, which may or may not explains the heterogeneity of non-specific cLBP. Using machine learning (ML) and data-driven approaches in a large set of metrics is well suited for discovering hidden patterns of variables (sub-grouping factors) and dividing subjects into homogeneous groups with unique characteristics (sub-groups). A recent work by Tagliaferri et all. (25) demonstrated that subgroups of cLBP patients can be discovered respect to a pain-free cohort using a combination of variable selection and clustering methods.

This work presents a machine learning (ML) and data-driven systems approach to learn subgrouping factors and patient subgroups for individuals admitted to a back pain specialty clinic - the UCSF Integrated Spine Service (ISS) between 2018 and 2020 - and its association with outcomes depending on non-standardized, patient-centric intervention patterns. We use different unsupervised ML techniques to discover multidimensional associations of variables available at clinical presentation, and homogeneous patient subgroups considering different data domains (e.g., demographics, pain characteristics, comorbidities). We compare SBT stratification with our ML-based process to determine further informative clinical features meaningful for patient subgrouping and prognosis.

## Methods

### Cohort identification and data extraction

ISS is a multidisciplinary program for improving the quality of care delivered to patients with spinal (cervical, thoracic, or lumbar) chronic pain. Patients are scheduled for back-to-back appointments with a physical therapist and a physician, who formulate a jointed treatment plan based on the principles of pain neuroscience education with the emphasis on self-care strategies, active rehabilitation, and non-interventional treatments. SBT is administered to all patients at baseline. There are no standardized treatment pathways, but patients who fall into the high-risk subgroup are discussed at monthly multidisciplinary case conferences so that the patient progress is closely monitored, and treatment is adjusted as indicated.

All adult patients diagnosed with cLBP and referred to ISS between 2018 and 2020 who completed baseline PROMIS-10 global health questionnaire and baseline SBT were included. The exclusion criteria for this study were consistent with the ISS exclusion criteria: presence of cancer, active spine infection, neurological deficits urgently needing surgery. Figure 1 provides a flow diagram of the included patients in the analysis. Ethics committee/IRB of the University of California San Francisco gave ethical approval for this work.

**Figure 1.**
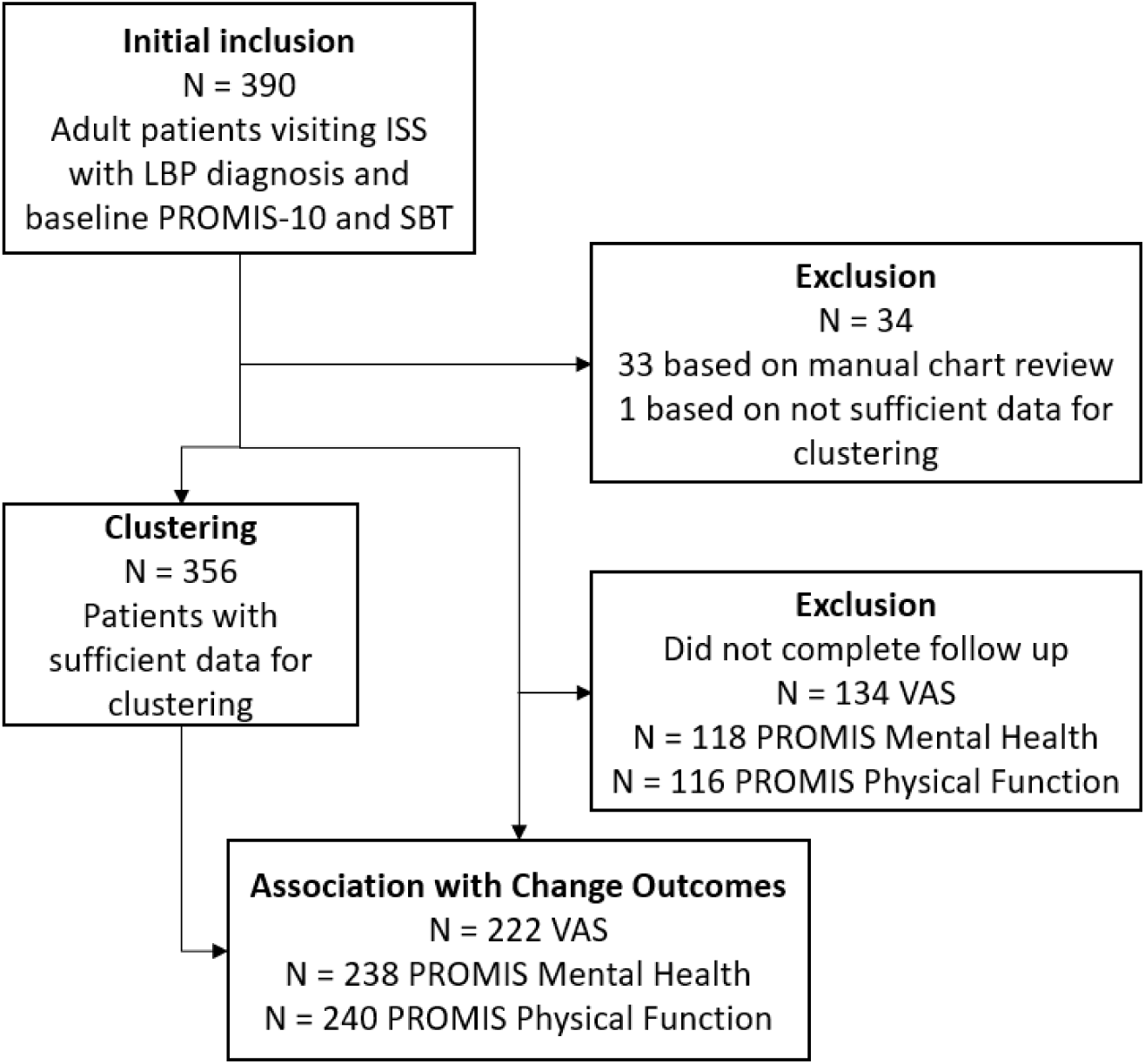
Flow diagram of dataset and analysis

### Data measurement

The VAS and PROMIS-10 global health questionnaire were administered at baseline. As part of a quality improvement project, a one-time follow up survey was conducted via phone or email in August 2020. Similarly, clinical notes of individual patients were reviewed at initial visit to characterize pain intensity (VAS), duration of pain, other pain locations, and presence of clinician diagnosed weakness. In addition, we obtained all other features available within the electronic medical record (EMR) at time of referral. This included using ICD-10 codes available in the chart to assess the patient’s Charlson Comorbidity Index (26), presence of overlapping pain conditions (COPCs) (13), prescription of medications including anti-inflammatory, analgesic, opioids, anti- convulsants, and glucocorticoids. Similarly, we tabulated utilization of imaging, emergency department visits, nerve tests, physical therapy visits, acupuncture, hospitalization, injections, and surgeries in the 6 months prior to the baseline visit, and from baseline visit and PROMIS-10 follow up time.

### Data pre-processing

Before conducting unsupervised learning, we performed the following data pre-processing steps: 1) data re-coding, 2) variable filtering, 3) missing value analysis and imputation. We first re- coded categorical variables using dummy variables or one-hot encoding including a variable for each level and unknown or missing cases (e.g., sex.male, sex.female, sex.unknown). Ordinal and continuous variables were used in their original form. Then, to reduce uninformative variables, we filtered out variables with near-zero variance (variance cutoff of 0.02).

#### Missing value analysis

The built dataset contains missing values (Supplementary Fig. 7), a characteristic of biomedical research and real-world data. We proceeded with data imputation through multiple imputation by chain equation (MICE) using the R mice package (27). We performed 100 multiple imputations to obtain 100 complete datasets through predicting mean matching. The complete datasets were then aggregated before unsupervised learning by the median value in quantitative variables and the mode in qualitative variables. Visual inspection of the distribution of the imputed vs. non-imputed values for each imputed variable revealed similar distributions (Supplementary Fig. 7)

### Unsupervised learning

#### Non-linear PCA (NLPCA)

We performed NLPCA by means of optimal scaling and alternating least squares implemented in the PRINCALS algorithm in the Gifi R package, which solves for the Gifi loss function (28). We chose this method because it allows for principal component (PC) extraction of mixed data while accounting for non-linearity in the variables through optimal scaling quantification or transformations. The transformations were considered as follow: categorical variables were not ordinally restricted, which allows for non-monotonic quantification based on the proportion of each level in the variable. Ordinal and continuous variables were ordinally restricted. Continuous and ordinal variables were transformed through a b-spline of degree 2 and 3 knots placed on the tertials (continuous) or the categories (ordinal) of the datapoints allowing for a non-linear transformation. The number of selected PC or dimensions was determined using the Kaiser (eigenvalue > 1) and elbow-in-Scree-plot criterions, with a final selection of the first 25 PCs. Examining the loading patterns, we observed that each PC is loaded by several variables with generally low loadings and each variable loads into several PCs. To simplify the PC solution and increase interpretability of the resulted dimensions, we performed varimax rotation of the loadings of the selected PCs (29). Varimax rotation was chosen since it preserves the distance between observations, increasing interpretability of the latent low dimensional coordinate system without affecting the internal structure of the dataset (i.e., the distance between subjects). The rotated latent dimensions were described by expert clinicians and researchers based on the variables with higher loadings (|loadings|>0.4). The rotated scores were calculated by applying (i.e., dot product) the resulting rotation matrix by the NLPCA scores of the 25 first PCs. The Median Hoffman index of complexity (30) for the unrotated original solution was 7.9 (the higher the value, the more complex the solution, with the maximum being equal to the number of total variables), indicative of high level of complexity in the latent solution (i.e., each PC is composed of several low to middle loadings and only captures a small portion of the variance), which made the interpretability more challenging. The new rotated dimensions had lower complexity than the original PCs (Hoffman index median = 1.89) with the consequent simplification of the loadings and increase interpretability.

#### Probabilistic-based clustering

Clustering was performed by modeling the rotated latent space through a mixture of gaussian distributions. Mixture models have previously shown good results for estimating densities and deriving clusters of patients in complex heterogeneous diseases (31). The models are parametrized by the mean vector of each cluster, and the covariance matrix of each cluster, and solved through the expectation maximization algorithm (32,33). The number of clusters (a.k.a., mixture components), and the different structural constrains that can be imposed to the covariance matrices are specified. Since our goal was to perform an unsupervised description of the structure of the data and we did not have a prior knowledge of the number of mixtures or the form of their covariance matrices, we performed a linear search by specifying a range of k= {1, …, 10} mixtures and different covariance structures. The limit was set to 10 after an exploratory analysis suggesting that more than 3-6 clusters were unlikely. To fit the models, we used the mclust R package (33), and chosen the covariance parametrization among the mclust implemented models m = {EII, VII, EEI, EVI, VEI, VVI, EEE, VEE, EVE, VVE, EEV, VEV, EVV, VVV}. These are defined by the cluster orientation (respect to coordinate system), volume (whether all cluster have a similar number of patients per cluster), and shape (whether all clusters have approximately the same multidimensional shape). We refer the reader to mclust documentation and further material (33) for details. For model selection, we first fitted all the models in m through k and used the Bayes Information Criteria (BIC) to compare models. Then we selected the top 3 performing models m = {VVI, VII, VEI} for further evaluation. Since selecting models that have close BIC can be problematic, we studied the stability of the model by leave-one-out (LOO) resampling. For each of the m models, k mixtures and n patients, a model was fitted on n-1 patients. The BIC and the log-likelihood of the patient left out (LOO- loglikelihood) for the n resulting models were calculated. For each m and k combination, the mean and 95% confidence interval of the BIC and LOO-loglikelihood were computed.

The data were best fitted by a 3-class model with VVI covariance structure based on model fit parameters and leave-one-out cross-validation (Supplementary Fig. 2). Increasing model complexity (i.e., number of classes) beyond three classes did not improve the model performance as it did not capture any additional underlying observations (Supplementary Fig. 2a-c), reinforcing the 3-VVI class model to be the best fit for our data.

#### Phenotypic characterization of clusters

To characterize the differences between clusters, a one-way ANOVA with phenotyping factor scores as response variable and cluster membership as predictor was used for each phenotyping factor. When the differences between clusters was significant (p < 0.05), a pairwise estimated marginal means contrast between clusters was performed (Supplementary Table 3).

### Analysis of outcome associations

To analyze the association between outcomes (delta PROMIS Physical Function, delta PROMIS Mental health, and delta VAS), we used linear models (ordinary least squares) with the outcomes as response variable. An F statistic test and ANOVA table were performed to determine the contribution and p value of the terms in the model. When multiple predictors were incorporated in the model, a stepwise forward and backward model selection procedure was used for selecting a parsimonious model based on Akaike Information Criteria (AIC) using R *step* function. P values were adjusted for the false discovery rate (Benjamini-Hochberg) and an adjusted p value < 0.05 was considered significant.

## Results

## Cohort

A total of 356 patients were included in the clustering analysis (Fig.1, Table 1). The median age was 61 (IQR: 48 - 72), 62% of the patients were females, and they exhibited different durations of pain, with 20% experiencing pain for 0-3 months and 41% for five years or more. The SBT categorized patients into Low (29%), Medium (42%), or High (29%) risk. The median PROMIS Mental Health standard score was 44 (IQR: 39 – 51), and the median PROMIS Physical Function standard score was 40 (IQR: 35 – 45). The median VAS was 7 (IQR: 5.75 – 8). Follow-up outcomes were obtained for VAS, PROMIS Mental Health, and PROMIS Physical Function. The median time for VAS follow-up was 56 days (IQR: 48 – 70), and for PROMIS was 529 days (IQR: 317 – 700). The distribution of delta values from the follow-up to baseline are shown in Figure 2.

**Table 1.** Patient characteristics at clinic presentation

|  | **N = 356<sup>†</sup>** |
| --- | --- |
| Age | 61 (48, 72) |
| BMI (kg/m<sup>2</sup>) | 26.6 (23.5, 30.4) |
| Unknown | 7 |
| Female | 222 (62%) |
| Insurance type |  |
| Private | 160 (45%) |
| Medical | 66 (19%) |
| Medicare | 130 (37%) |
| Race |  |
| White/Caucasian | 173 (49%) |
| Black/African American | 51 (14%) |
| Asian | 81 (23%) |
| Others and unknown | 51 (14%) |
| Smoking |  |
| Never Smoker | 208 (59%) |
| Former Smoker | 130 (37%) |
| Current Some Day Smoker | 8 (2.3%) |
| Current Every day Smoker | 9 (2.5%) |
| Unknown | 1 |
| Pain Duration |  |
| 0-3 months | 64 (20%) |
| 3-6 months | 24 (7.6%) |
| 6-12 months | 33 (10%) |
| 12-24 months | 22 (6.9%) |
| 24-36 months | 14 (4.4%) |
| 36-48 months | 13 (4.1%) |
| 48-60 months | 17 (5.4%) |
| 5 years or more | 130 (41%) |
| Unknown | 39 |
| StartBack risk assessment |  |
| Low Risk | 102 (29%) |
| Medium Risk | 145 (42%) |
| High Risk | 100 (29%) |
| Unknown | 9 |
| PROMIS Mental Health score (higher = better) | 44 (39, 51) |
| Unknown | 15 |
| PROMIS Physical Function score (higher = better) | 40 (35, 45) |
| Unknown | 10 |
| VAS score (higher = worse pain) | 7.00 (5.75, 8.00) |
| Unknown | 20 |
| CCI | 1.00 (0.00, 2.00) |

**Table 1.** Patient characteristics at clinic presentation
|  | **N = 356<sup>1</sup>** |
| --- | --- |
| <sup>1</sup> Median (IQR); n (%). BMI: body mass index; PROMIS: Patient-Reported Outcomes Measurement Information System; VAS: Visual Analog Scale; CCI: Charlson Comorbidity Index. |  |

**Figure 2.**
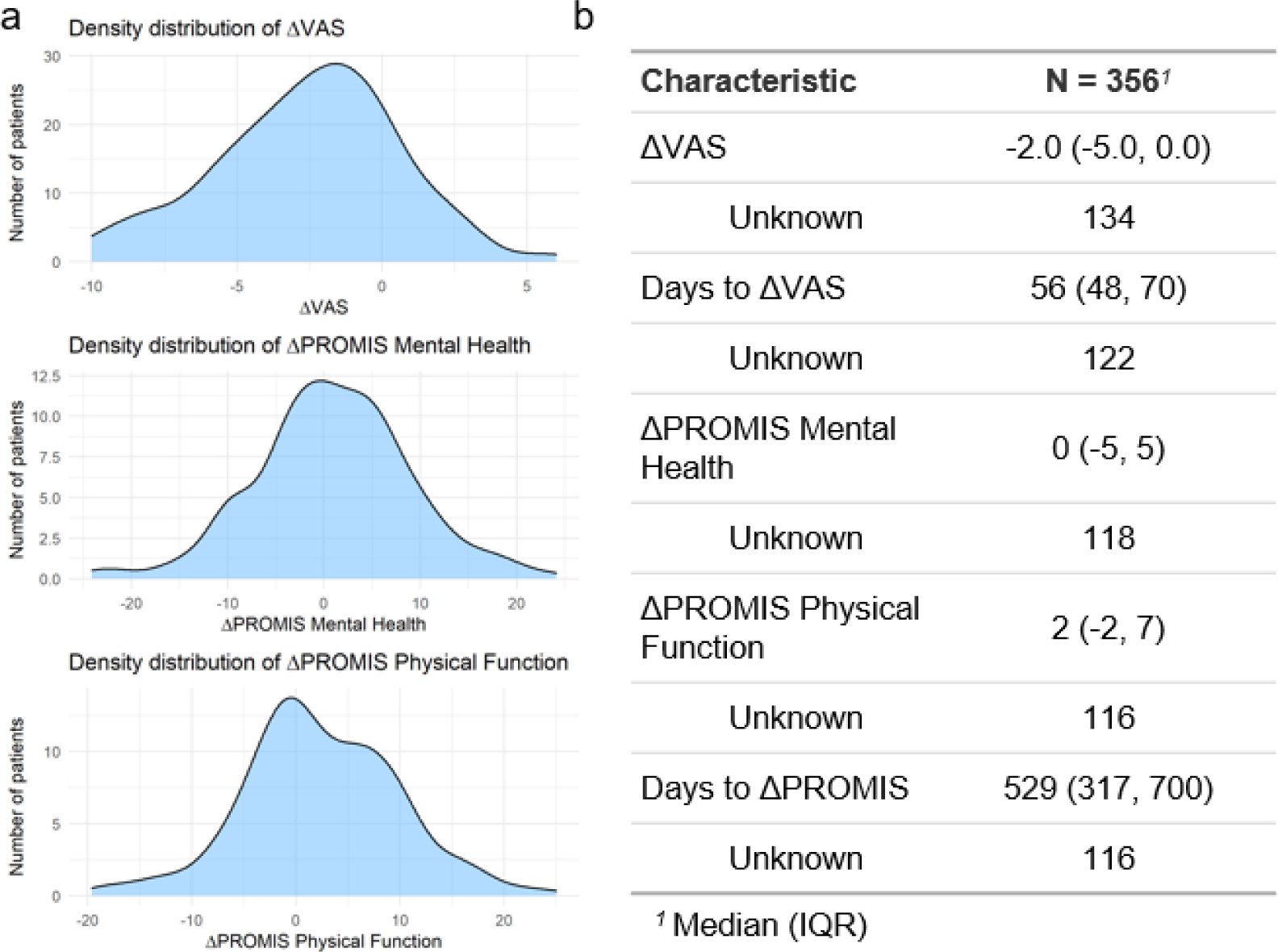
Delta values from the follow-up to the baseline. The estimated frequency distribution of the delta values for the three outcomes are shown in **a**. The median (IQR) delta value and the median (IQR) of days for which the delta was measured are shown in **b.** IQR: Interquartile range.

## Patient phenotypes

We used a two-step ML workflow to derive patient phenotypes with data for the 6 months before first visit, referred to as pre-presentation (e.g., comorbidities, medications and procedures) and data at the first visit, referred to as data at-presentation (e.g., demographics, pain characteristics). We first grouped variables by common (latent) phenotypic factors using Non-Linear Principal Component Analysis (NLPCA) (Supplementary Fig. 1), effectively reducing the dimensionality of the data. NLPCA was used as it allows for mixed variable types (continuous, nominal, and ordinal) through non-linear transformations of the data (28,34). Then we clustered patients based on their individual scores in the NLPCA, which we named phenotyping factors scores (PFS).

We retained the first 25 principal components (PCs) based on the Kaiser and elbow criteria (see methods), which represent the PCs that capture variability above what is expected for a single variable. The 25 PCs explained an accumulated total variance of 69.4%. The PCs were rotated using varimax rotation to increase their interpretability, and the resulting latent dimensions were regarded as the relevant phenotypic factors interpreted by expert clinicians (Supplementary Table 1). Overall, the complex multidimensional structure of the pre-presentation and at-presentation data indicates cohort heterogeneity and suggests several factors that determine patient phenotypes. Next, we derived clusters of patients using probability-based clustering through gaussian mixture models (GMM) of PFS, resulting in three clusters (Fig. 3a; Supplementary Fig. 2). Each patient was assigned to a given cluster by their maximal posterior probability resulting in clusters of 96, 126, and 134 patients, respectively (Table 2). Since SBT is used for stratifying patient risk of future disabling low back pain, we compared whether the 3 clusters reassemble SBT groups at the first ISS visit. The Adjusted Rand Index, a measure of agreement from 0 (no agreement) to 1 (exact partitions), was 0.021. This indicate that the stratification generated by the three clusters is different than the one given by the SBT alone. Note that these are not necessarily independent as SBT was included in the ML procedure (Chi-square independence test of SBT vs clusters partitions p < 0.001), but it suggests that adding more information beyond SBT for partitioning patients generates different subgroups.

**Figure 3.**
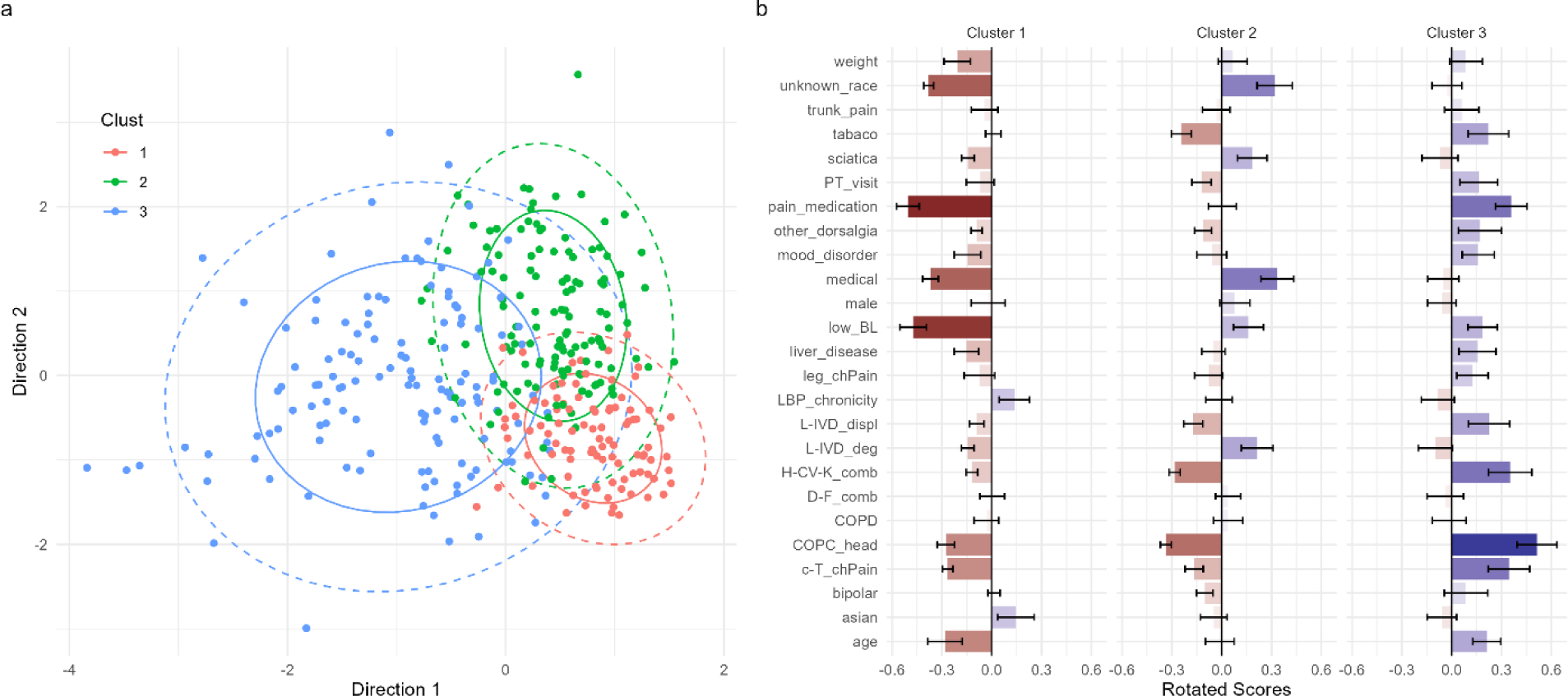
Clustering of the phenotyping factors scores revealed 3 clusters (Supplementary Fig. 2). The density estimate for each gaussian mixture (i.e., cluster) can be represented in a scatterplot of the 2 most discriminant directions from the phenotyping factors rotated scores (**a**). Each dot represents a subject colored by the most probable assigned cluster. Solid ellipsoid represents the bivariate standard deviation, and the dashed ellipsoid represents the 95% bivariate confidence interval for each cluster. The closer two subjects are, the more similar they are in their multidimensional phenotype. The average ± SE phenotyping factor scores for each factor and cluster are shown in (**b**). The more positive the score is the higher the factor is represented in the cluster, and vice-versa for negative scores (e.g., positive score for age represents older than average, negative score represents younger than average). Values of 0 represents the average score of the entire cohort.

**Table 2.**
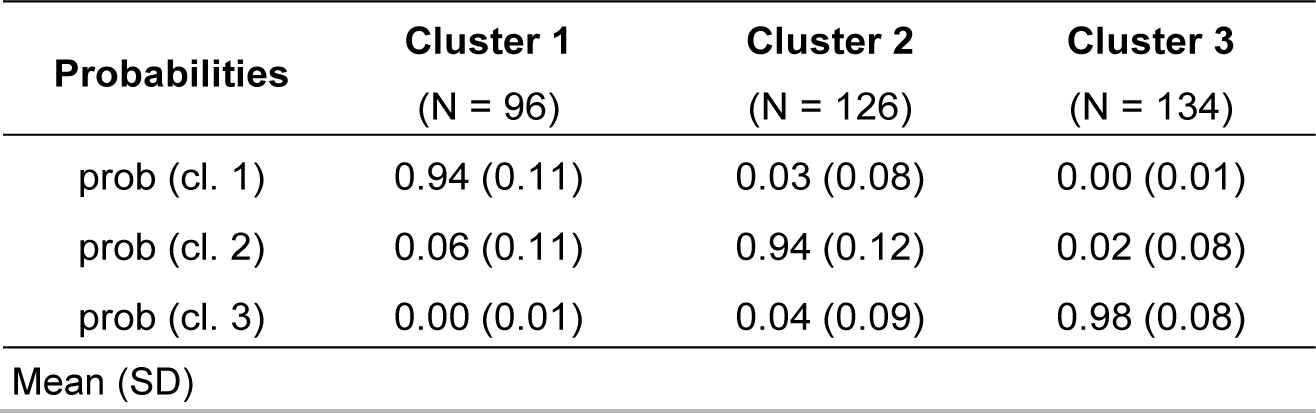
Cluster classification probabilities of presentation data

The three clusters were described by their PFS, providing the characteristics defining a given patient phenotype (Fig. 3b). PFS value closest to 0 means that patients locate around the factor average of all patients; positive values for a given phenotypic factor mean that patients in that cluster have a higher representation of that phenotypic factor than the average, and vice versa for negative scores. Cluster 1 is generally characterized by a younger population, low pain medication prescription, high function with low pain at presentation, a high proportion of patients with private insurance, and a low number of overlapping pain conditions. Cluster 2 is characterized by a high proportion of patients with Medicaid insurance, enriched with patients of other or unknown race (27.8% of the group), presenting with lower number of comorbidities and chronic overlapping pain conditions (COPCs), especially head pain, and low tobacco use. Cluster 3 captures older participants characterized by high COPC and head pain, comorbidities, and pain medication use pre-visit to ISS. We then compared the differences between clusters for each of the phenotypic factors (Supplementary Tables 2 and 3). The age factor was significantly different between clusters 1 and 3, with a gradual increase between clusters 1 (negative), 2 (around 0), and 3 (positive). Cluster 3 was significantly higher in COPC and head pain, cervico-thoracic chronic pain, heart, cerebrovascular, and kidney comorbidities than clusters 2 and 1, with no differences between the latter two. Regarding baseline pain and function, Cluster 1 presented significantly lower PFS than clusters 2 and 3. The proportion of individuals of other/unknown races was lowest in cluster 1, average in cluster 3, and the highest in cluster 2. Finally, tobacco use was reported higher by the individuals in cluster 3 than those in cluster 2. The complete list of comparisons can be found in Supplementary Tables 2 and 3.

Using a similar NLPCA workflow we obtained a small set of variables that captures different profiles of interventions administered in the population (Table 3; Supplementary Fig. 3). The three presentation clusters were mainly differentiated on whether they received pain medication (p<0.01). On average, patients in cluster 1 received less medication after the first ISS visit than patients in cluster 2 and 3, while cluster 3 had the highest average in pain medication. Accordingly, 69% of patients in cluster 3 were prescribed opioids, with gradual reduction to 48% and 33% of patients with opioid prescription in clusters 2 and 1, respectively (Chi-squared p < 0.001). A similar pattern was observed for anti-inflammatory analgesics (Chi-squared p <0.001; cluster 1 31%, cluster 2 49%, cluster 3 60%) (Supplementary Fig. 4)

**Table 3.** Mean values of Intervention factor scores per cluster (univariable analysis)

| Intervention factor | Cluster 1<br>(N = 96) | Cluster 2<br>(N = 126) | Cluster 3<br>(N = 134) | P value* |
| --- | --- | --- | --- | --- |
| Antidepressant | -0.10 (0.84) | 0.07 (0.97) | 0.01 (1.13) | 0.44 |
| In-patient hospitalization | -0.03 (0.79) | -0.02 (0.99) | 0.04 (1.15) | 0.81 |
| Pain medication | -0.47 (0.94) | -0.01 (0.93) | 0.34 (0.98) | **<0.001** |
| ED visit | -0.05 (0.69) | 0.01 (1.08) | 0.02 (1.11) | 0.84 |
| Surgery | 0.11 (0.85) | -0.07 (0.98) | -0.01 (1.12) | 0.42 |
| Acupuncture | -0.01 (0.78) | 0.07 (1.18) | -0.06 (0.96) | 0.54 |
| PT visit | 0.10 (1.03) | -0.01 (0.96) | -0.06 (1.02) | 0.52 |

Mean (SD). \*P value of one-way ANOVA

## Phenotypes associations with outcomes

Subjects in either phenotype showed a significant average reduction in delta VAS (overall change -2.57 [-2.99, -2.15], p < 0.001), while subject in phenotype 2 showed a significant average increase in PROMIS Mental Health scores (1.78 [0.037, 3.52], 0.045), and PROMIS Physical Function scores (3.2 [1.585, 4.81], p < 0.001) with respect to the null hypothesis of no change. Phenotype 3 presented a significant average increase in PROMIS Physical Function scores (2.41 [0.86, 3.95], p = 0.002) (Fig. 4a-c).

**Figure 4.**
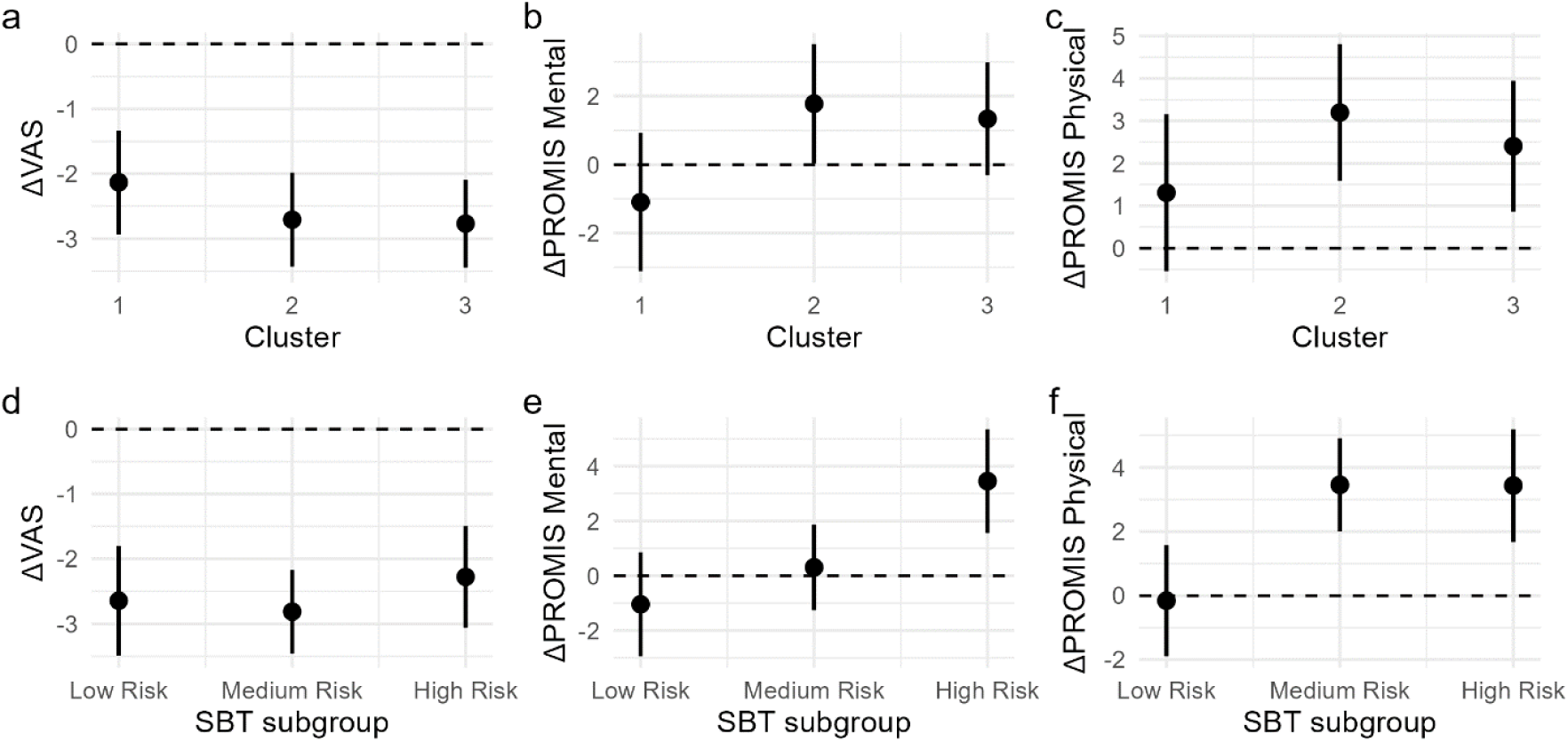
Delta outcomes differences per subgrouping method. No statistical significant differences between cluster membership in the prediction of outcome were observed (**a-c**), while there are significant differences with SBT subgrouping(**d-f**) in change in PROMIS Mental Health (**e**) and PROMIS Physical function (**f**). Graph represents the expected marginal mean (dot) and 95% confidence interval (error bar) of the analysis model (Table 4). Dashed line represents no change between follow-up and baseline.

No statistical differences were found when comparing the three phenotypes. To reduce the effect of uncertainty on cluster allocation, we performed the same analysis considering only patients with high certainty of cluster membership (posterior probability of 80% or higher). This increases the effect sizes although still not reaching significance when comparing among phenotypes (Supplementary Table 4). This is in contraposition of SBT (Fig. 4d-f), where we observed a significant association of changes in PROMIS Mental Health (p < 0.01) and PROMIS Physical Function (p < 0.01) scores with SBT-defined subgroups.

## Phenotyping factors associations with phenotypes and outcomes

Next, we sought to determine the minimal number of phenotyping factors that could predict outcomes by using multivariate regression with model selection. Table 4 show a summary of the significant factors, with complete tables in the supplement (Supplementary Tables 5, 6, 7).

**Table 4.** ANOVA and marginal effects of significant phenotyping factors associated to outcomes (multivariable analysis)

| **Delta PROMIS Physical (n = 240)** |  |  |  |  |
| --- | --- | --- | --- | --- |
| **Phenotyping factor** | **F value** | **Adj. P value** | **Coeff.** | **Lower, Upper CL** |
| COPC_head | 10.46 | 0.01 | -1.34 | -2.17, -0.49 |
| leg_chPain | 6.73 | 0.04 | 0.97 | 0.11, 1.83 |
| low_BL | 32.40 | p<0.01 | 2.37 | 1.48, 3.26 |
| COPD | 6.63 | 0.04 | -1.13 | -1.99, -0.26 |
| asian:cluster | 8.10 | p<0.01 |  |  |
| asian:cluster 1 |  |  | -2.08 | -3.52, -0.63 |
| asian:cluster 2 |  |  | 1.62 | 0.1, 3.136 |
| asian:cluster 3 |  |  | -2.05 | -3.35, -0.73 |
| **Delta PROMIS Mental (n = 238)** |  |  |  |  |
| **Phenotyping factor** | **F value** | **Adj. P value** | **Coeff.** | **Lower, Upper CL** |
| COPC_head | 14.25 | p<0.01 | -1.63 | -2.54, -0.72 |
| low_BL | 30.57 | p<0.01 | 2.75 | 1.76, 3.74 |
| **Delta VAS (n = 222)** |  |  |  |  |
| **Phenotyping factor** | **F value** | **Adj. P value** | **Coeff.** | **Lower, Upper CL** |
| medical | 7.84 | 0.04 | 0.43 | 0.04, 0.83 |
| L-IVD_displ | 6.23 | 0.04 | 0.39 | 0.003, 0.77 |
| Asian | 6.29 | 0.04 | 0.47 | 0.07, 0.88 |
| PT_visit:cluster | 4.58 | 0.04 |  |  |
| PT_visit:cluster 1 |  |  | 1.22 | 0.27, 2.17 |
| PT_visit:cluster 2 |  |  | 0.65 | -0.35, 1.667 |
| PT_visit:cluster 3 |  |  | 0.04 | -0.41, 0.5 |
| L-IVD_deg:cluster | 5.20 | 0.04 |  |  |
| L-IVD_deg:cluster 1 |  |  | -3.69 | -5.72, -1.66 |
| L-IVD_deg:cluster 2 |  |  | -0.26 | -0.83, 0.3 |
| L-IVD_deg:cluster 3 |  |  | -0.55 | -1.06, -0.05 |
| *Note: Only significant predictors are shown, see full table in supplementary Table 5, 6, and 7* |  |  |  |  |

The final parsimonious model for delta PROMIS Physical Function explained 37.4% of the variance (Supplementary Table 5). An increase in Physical Function was associated with higher values of the “Low baseline function” factor (p < 0.01) and “Leg pain” factor (p = 0.04), and lower values of “COPC and head pain” (p = 0.01), and “COPD” (p = 0.04). “Asian vs. White” phenotyping factor presented a significant interaction with the clusters in association with Physical Function (p < 0.01). The interaction shows that being Asian was associated with better improvement of Physical Function in phenotype 2, while it was associated with less improvement in phenotypes 1 and 3 (Supplementary Fig. 4). In the case of delta PROMIS Mental Health, the final model explained 33.02% of the variance (Supplementary Table 5). Like physical function, increase in mental health function was related to higher values of “Low baseline function” factor, and lower values of “COPC and head pain”. No interactions between factors and phenotypes were found significant. Finally, reduction in pain (delta VAS, Supplementary Table 6) was associated with lower values of the factors “Medicaid insurance”, “Lumbar IVD displacement” and “Asian vs. White”. Differences between phenotypes on the association to delta VAS (significant interaction) were found for the factor “PT visits for LBP” (p = 0.04) and “Lumbar IVD degenerations” (p = 0.04). A post-hoc of these interactions showed that patients with positive values in the “PT visits for LBP” factor (more likely to have visited with a PT before ISS) are less likely to reduce pain if they are of phenotype 1. This relationship is not observed for phenotypes 2 and 3. On the other hand, patients in phenotype 1 that presents to the ISS with Lumbar IVD degeneration are more likely to reduce their pain after ISS visits than those patients with no Lumbar IVD degeneration (Supplementary Fig. 5).

## Intervention factors associations with phenotypes and outcomes

Lastly, we studied the association of the interventional factors with phenotypes and outcomes. An increase in Physical Function was associated with higher values of the intervention factor “ED visits” (p = 0.01), with no other factors nor interactions with phenotypes being significant (Supplementary Table 8). The final delta PROMIS Mental Health model explained only 6.35% of the variance, and no significant associations were found for the included intervention factors (Supplementary Table 9). Similarly, no significant factors were associated with delta VAS, with a final model explaining 12.4% of the variance (Supplementary Table 10).

## Discussion

In this study we evaluated patient heterogeneity in a cohort of patients treated in a cLBP specialty clinic. We found several phenotyping factors associated with patient outcome and 3 distinctive patient subtypes or clinical phenotypes. Specifically, using data-driven unsupervised dimensionally reduction and probabilistic-based clustering we identified 25 independent factors capturing different patient characteristics at presentation to ISS, 7 independent factors capturing interventions undertaken post-ISS presentation, and 3 patient sub-populations with different characteristics.

### Heterogeneity of cLBP

Our findings reveal a highly complex cohort, consistent with the notion of heterogeneity among cLBP patients. Despite dimensionality reduction 25 principal components, accounting for an accumulated variance of 69.4%, were considered informative. This contrasts with previous studies where only 3 PCs were retained (35). Our results indicate that there is redundancy in the original set of variables, but also that the true dimensionality of the data is high, with at least 25 meaningful dimensions and several interrelations between variables (complex PC composition). Tagliaferri et all. (25) used a univariate approach for initial variable selection before clustering cLBP patients. This has the risk of discarding variables from the analysis that are informative for clustering when combined with other variables. In contrast, our results suggest that there are emergent characteristics in cLBP that can be measured from the combination of observed variables, and those could be meaningful for patient phenotyping. We captured expert opinion on the definition of these emergent properties in order to clarify their clinical meaning. Some of these characteristics have established associations with cLBP: chronicity, chronic other site pain (COPC and head pain, cervico-thoracic chronic pain, leg chronic pain, presence of sciatica), diabetes and fibromyalgia comorbidities, pain medication use, presence of mood disorder/bipolar disorder, tobacco use. Other important characteristics identified in this work had only weak or no associations with cLBP outcomes in prior studies such as higher weight, older age, male sex, disc degeneration, and race (particularly Asian). Thus, many of the findings from data-driven unsupervised learning are consistent with prior knowledge, while others provide novel insights. The latter may reflect clinically important association that can drive new directions for determining patient subgroups and prognostic predictors.

We identified 3 phenotypes distinct of the three subgroups determined by SBT. Cluster 1 consists of those individuals with characteristics found to be protective of chronic pain: younger age, low pain medication prescription, high function, good insurance access, and low overlapping pain conditions. Individuals in Cluster 3 associate with older age and present with higher incidence of COPC and head pain, comorbidities, and pain medication use. Cluster 2 is an intermediate group with an enrichment of patients of the other or unknown race. It is not clear whether “other/unknown” race encompasses groups historically associated with disparate outcomes. Thus, further exploration is warranted. Use of pain medication seems to be the single intervention factor that significantly differs among the 3 clusters, with opioid prescription being the biggest difference. Cluster 1, was associated with the least pain medication prescription, including both opioids and anti-inflammatory analgesics. These differences follow the same pattern previously ISS visit, with the tendency of pain medication prescription continuing for those patients who are already on medication after ISS visits. It is possible that differences in pain medication prescription underly the different phenotypes. For example, long-term pain medication use have significant changes in response to pain such as hyperalgesia (opioid-induced hyperalgesia)(36), which could lead to effects that might be captured in the discriminatory variables between clusters. A future time-based analysis of causal effects with a more granular time event data might help better study how pain medication may be highly linked to separations in cLBP subgroups and whether prolonged pain medication may play a role in chronification of pain (37).

Other measurements beside what is included in this work could be also important for defining cLBP subgroups. We have previously shown that biomechanical metrics automatically extracted using markerless motion capture during the sit-to-stand task can discriminate patient subgroups and relate to pain and disability (38). In addition, previous clustering work have noted psychosocial variables to be important for separating cLBP into distinct sub-groups. Bäckryd et al., found 4 clusters largely based on psychosocial and pain related questionnaires (35), with subgroups characterized mostly for their differences in psychological strain levels, social support, and pain characteristics such as intensity and duration. Their subgroup presenting moderate pain intensity, overall better health and low psychological comorbidities presents similarities with our cluster 1 with lower VAS and SBT at baseline, and higher values of PROMIS-10 mental health. On the other side, they found a subgroup associated with lower health, high pain and high psychological strain, which share similarities with the phenotype described by our cluster 3. Tagliaferri et all. (25) found 5 subgroups of cLBP patients, mostly divided with reference to depressive symptoms and social isolation. Our data did not contain detailed measurements of psychosocial factors, which could have limited partitioning subgroups into further clusters. In contrast, our dataset contained a more detailed clinical description using data from medical records for months previous to ISS first visit. For example, prescribed pain medications are important discriminatory factors between clusters in the present study. Future work directed to cLBP subgrouping should consider a more comprehensive dataset including detailed descriptions from different domains such as demographics, medical records, psychosocial, pain processing, and biomechanics and physical function (5).

### Phenotypes associations with outcomes

Phenotypes did not statistically differ among them related to changes in physical, mental or pain scores. Nonetheless, clusters 2 and 3 showed average significant improvement in physical function, while cluster 1 did not. Since patients in Cluster 1 are already characterized by relatively higher function and younger age as compared to those in Clusters 2 and 3, it is possible that there is a ceiling effect limiting the observation of physical function improvement in those individuals.

The high heterogeneity of the cohort may require a higher number of subjects to resolve more meaningful patient subgroups using unsupervised methods. Indeed, the post-hoc diagnostic of the clustering partitions suggests that more clusters are possible, but there is not enough data to estimate them reliably. Considering only patients with high certainty of cluster membership increased the effect sizes of the differences between clusters regarding changes in outcome, although still without reaching statistical significance. This suggests that with a large enough dataset, we might be able to achieve clustering that can accurately predict outcomes. Furthermore, our approach is unsupervised, meaning outcome information is not considered when forming the subgroups. Stratification strategies with respect to outcomes using semi-supervised clustering could be an alternative and informative way to discover subgroups informing cLBP prognostics.

The phenotyping factors found through NLPCA had associations with outcomes. An increase in Physical and Mental functions was associated with “low baseline function” factor, mainly constituted by a combination of PROMIS Physical, PROMIS Mental and VAS scores, and SBT subgroups at presentation. Patients presenting to the ISS with a generally low function are those who improve the most. Although we cannot discard a regression to the mean effect between baseline and follow-up functional metrics, these results suggests a strategy for patient stratification and prognosis by augmenting SBT subgrouping with PROMIS-10 and VAS scores at baseline. Only considering SBT subgrouping explains 4%, and 3.9% of the change in PROMIS Physical and Mental scores from baseline to follow up, respectively. This value significantly increases to 9% (LRT p < 0.001) and 8.5% (LRT p = 0.001), respectively, when considering the “low baseline function” factor together. Future work should be directed at finding the best combination of SBT and baseline outcomes for creating a patient stratification and prognostic metric. Low values of the “chronic overlapping pain conditions and head pain” factor was related to higher improvement in physical and mental scores after admission to the ISS. COPCs captures the comorbidity of distinct pain conditions, such as headache, low back pain, fibromyalgia, temporomandibular disorders, and irritable bowel syndrome, but share common comorbidities and risk factors such as sex, increased pain sensitivity, and genetic variants (39,40). The presence of COPCs has been associated with worse outcomes for the treatment of other pain conditions, such as chronic migraines (41). Although we could not fine evidence of cluster association with outcomes, cluster 3 was characterized with high values in COPCs and head pain. Since COPCs acompasses other symptoms beyond pain such as fatigue, sleep impairment and physical and mental dysfunction (40), it is possible that the presence of COPCs is a proxy for underlying biological and psychosocial mechanistic determinants of cluster 3 phenotype. A state of increased pain sensitivity or pain amplification might be higher when some pain conditions develop such as temporomandibular disorder (42), and potentially other COPCs (40). Peripheral and central nervous system dysregulation and sensitization underlying pain amplification is likely to also contribute to other symptoms such as altered mood, and sensory, motor and autonomic dysfunction (40,43,44). Subjects in cluster 3 also show, on average, high incidence of heart, cerebrovascular and kidney comorbidities and the presence of mood disorders such as anxiety and depression. Although it is impossible to establish a causal connection from our analysis, it could be the case that cluster 3 capture patients with nervous system dysregulation and sensitization as their central pathophysiological mechanism. Further research is needed to link cluster 3 phenotype to central sensitization as potential common mechanisms of pain and other symptoms. If this hypothesis is true, it could have implications on treatment options for subjects showing cluster 3 phenotype. In addition, our findings suggest screening for COPCs at the time of referral to specialty care as additional stratifying/phenotyping factor.

Interestingly, we observed an interaction between the phenotyping factor that represented being Asian with the phenotype in relation with changes in physical function. Asian patients in cluster 2 had greater physical function improvement than those that are not Asian, while this relationship is not observed for clusters 1 and 3. In fact, Asians in cluster 2 presented with the highest recovery in physical function. Although we cannot fully explain these relationships with the data at hand, understanding this interaction provides the opportunity to find novel determinants of functional recovery related to patient race/demographics. Our observation also suggests that race cannot be taken at face value since complex interactions may be present when phenotyping cLBP. It will be important for future studies to further delve into potential cultural, genetic, or treatment-related factors that might explain these relationships and to explore if similar interactions exist with other racial or ethnic groups and how they may influence recovery in cLBP.

### Limitations

One limitation to our work is that the data comes from a single-center with a relatively low number of subjects included in the analysis. Furthermore, this is an observational study, and the possibility of spurious associations must also be considered. Future work should focus on validating our clustering model and phenotypes. The inclusion of more patient data from other specialty clinics will potentially increase the population heterogeneity, providing better substrate to find more homogeneous patient strata with different medical needs. Finally, other combination of ML methods might produce different subgroups, and future research should be directed on determining which methods produce more stable, generalizable and clinically useful results.

## Conclusions

Overall, we demonstrate that cLBP population heterogeneity is quantifiable and that ML analytical workflow can be used to explain, in part, the heterogeneity in relation to outcomes. Notably, considering a data-driven approach from multi-domain data produces different subgroups than SBT, and the addition of other functional metrics at baseline such as PROMIS and VAS increases the variance explained in outcomes.

## Data Availability

All data produced in the present study are available upon reasonable request to the authors

## Acknowledgments

Research reported in this publication was supported by the National Institute Of Arthritis And Musculoskeletal And Skin Diseases of the National Institutes of Health under Award Number U19AR076737. AS acknowledges funding from National Science Foundation Grant DMS- 2210206.

