## Supplementary figures and tables for "Unsupervised subgrouping of chronic low back pain patients treated in a specialty clinic"

**Supplementary material**

**Supplementary Table 1**: Salient variables (|loading|>0.4) in the varimax-rotated NLPCA solution

| **Phenotyping factor label (short name)** | **Salient variable** | **Salient varimax loadings** |
| --- | --- | --- |
| ***Higher weight (weight)*** | weight | 0.79 |
|  | BMI | 0.88 |
|  | DX_Overweight.Obesity | 0.71 |
| ***Being older (age)*** | age | 0.82 |
|  | Finance_Class_reduced.private | -0.69 |
|  | Finance_Class_reduced.medicare | 0.87 |
|  | CCI_score | 0.53 |
|  | DX_Hypertension | 0.52 |
| ***Being male (male)*** | weight | 0.41 |
|  | sex.male | 0.92 |
|  | sex.female | -0.91 |
|  | height_cm | 0.74 |
| ***COPC and head pain (COPC_head)*** | ChPain_headFace | 0.43 |
|  | COPC_count_preISS | 0.88 |
|  | DX_Migraine | 0.82 |
| ***Cervico-thoracic Chronic Pain (C-T_chPain)*** | ChPain_neckUpperBack | 0.48 |
|  | ChPain_rightUpper | 0.81 |
|  | ChPain_leftUpper | 0.78 |
| ***Pain medication use (pain_medication)*** | MED_lv1_Analgesic.Anty.inflammatory | 0.56 |
|  | MED_lv1_OPIOID | 0.76 |
|  | MED_lv1_Other | 0.46 |
|  | MED_lv1_Other.anticonvulsants | 0.69 |
|  | MED_lv1_Anty.depresant | 0.41 |
| ***Unknown race (unknown_race)*** | race_recoded.otherUnknown | 0.89 |
| ***Leg Chronic Pain (leg_chPain)*** | pain_down_leg | 0.68 |
|  | ChPain_rightLower | 0.69 |
|  | ChPain_leftLower | 0.67 |
| ***Liver disease (liver_disease)*** | DX_Liver.disease | 0.62 |
|  | PROC_LBP_Outpatient.visit | 0.41 |
| ***Medicaid vs private (Medicaid)*** | Finance_Class_reduced.private | -0.51 |
|  | Finance_Class_reduced.medicaid | 0.79 |
|  | race_recoded.black_AA | 0.43 |
| ***High pain low function baseline (low_BL)*** | StartBack_risk_BL | 0.8 |
|  | PROMIS_Mental_standardScore_BL | -0.72 |
|  | PROMIS_Physical_standardScore_BL | -0.79 |
|  | VAS_BL | 0.43 |
| ***Chronicity of LBP (LBP_chronicity)*** | Chronicity_dis | 0.68 |
|  | Primary.Back.Pain | 0.68 |
| ***Heart, cerborvascular and kindey comorbidities (H-CV-K_comb)*** | DX_Chronic.heart.failure | 0.68 |
|  | DX_Cerebral.vascular.disease | 0.51 |
|  | DX_Kidney.renal.disease | 0.72 |
| ***Presence of Mood disorder (mood_disorder)*** | DX_Anxiety.disorder | 0.7 |
|  | DX_Depression | 0.74 |
| ***Lumbar IVD displacement (L-IVD_displ)*** | DX_Other.intervertabral.disc.displacement..lumbar.region | 0.72 |
| ***Tabaco use (tabaco)*** | DX_Tobacco.nicotineuse | 0.75 |
| ***PT visit for LBP (PT_visit)*** | DX_Low.back.pain | 0.48 |
|  | PROC_LBP_Imaging | 0.47 |
|  | PROC_LBP_PT | 0.68 |
| ***Presence of Sciatica (sciatica)*** | DX_Sciactica..unspecified.side | 0.68 |
| ***Presence of Bipolar disorder (bipolar)*** | DX_Bipolar.disorder | 0.72 |
| ***Diabetes and fibromyalgia comorbidities***  ***(D-F_comb)*** | DX_Diabetes.type.II | 0.51 |
|  | DX_Fibromyalgia | 0.5 |
| ***Chronic pulmonary disease (COPD)*** | DX_Chronic.pulmonary.disease | 0.75 |
| ***Presence of other dorsalgia (other_dorsalgia)*** | DX_Other.dorsalgia | 0.78 |
|  | PROC_LBP_ED | 0.44 |
| ***Lumbar IVD degeneration (L-IVD_deg)*** | DX_Other.interveterbral.disc.degeneration..lumbar.region | 0.69 |
| ***Trunk pain (trunk_pain)*** | ChPain_trunk | 0.71 |
| ***Asian vs. White (asian)*** | race_recoded.white_caucasian | -0.59 |
|  | race_recoded.asian | 0.77 |
|  | Smoking_Status_recoded | -0.42 |

**Supplementary Table 2**: ANOVA of cluster differences for each phenotyping factor

| **Phenotyping factor** | **Df** | **Sum Sq** | **Mean Sq** | **F value** | **Adj p value** |
| --- | --- | --- | --- | --- | --- |
| age | 2.00 | 13.61 | 6.80 | 7.01 | **0.02** |
| asian | 2.00 | 2.80 | 1.40 | 1.40 | 1.00 |
| bipolar | 2.00 | 2.33 | 1.16 | 1.16 | 1.00 |
| c-T_chPain | 2.00 | 26.21 | 13.11 | 14.03 | **<0.01** |
| COPC_head | 2.00 | 56.91 | 28.45 | 33.58 | **<0.01** |
| COPD | 2.00 | 0.32 | 0.16 | 0.16 | 1.00 |
| D-F_comb | 2.00 | 0.39 | 0.19 | 0.19 | 1.00 |
| H-CV-K_comb | 2.00 | 28.12 | 14.06 | 15.14 | **<0.01** |
| L-IVD_deg | 2.00 | 9.08 | 4.54 | 4.62 | 0.16 |
| L-IVD_displ | 2.00 | 11.29 | 5.64 | 5.78 | **0.05** |
| LBP_chronicity | 2.00 | 2.75 | 1.37 | 1.37 | 1.00 |
| leg_chPain | 2.00 | 3.40 | 1.70 | 1.70 | 1.00 |
| liver_disease | 2.00 | 5.80 | 2.90 | 2.92 | 0.62 |
| low_BL | 2.00 | 29.39 | 14.69 | 15.88 | **<0.01** |
| male | 2.00 | 1.29 | 0.64 | 0.64 | 1.00 |
| medicaid | 2.00 | 27.55 | 13.78 | 14.81 | **<0.01** |
| mood_disorder | 2.00 | 5.93 | 2.96 | 2.99 | 0.62 |
| other_dorsalgia | 2.00 | 6.21 | 3.11 | 3.14 | 0.58 |
| pain_medication | 2.00 | 41.74 | 20.87 | 23.45 | **<0.01** |
| PT_visit | 2.00 | 5.86 | 2.93 | 2.96 | 0.62 |
| sciatica | 2.00 | 6.86 | 3.43 | 3.47 | 0.45 |
| tabaco | 2.00 | 14.06 | 7.03 | 7.26 | **0.01** |
| trunk_pain | 2.00 | 0.81 | 0.40 | 0.40 | 1.00 |
| unknown_race | 2.00 | 26.87 | 13.44 | 14.41 | **<0.01** |
| weight | 2.00 | 5.71 | 2.86 | 2.88 | 0.62 |

**Supplementary Table 3:** Pairwise marginal mean contrast between clusters for each phenotypic factor with mean cluster effect (ANOVA, Supplementary Table 2).

| **Contrast** | **Phenotyping factor** | **estimate** | **SE** | **t.ratio** | **p.value** |
| --- | --- | --- | --- | --- | --- |
| Cluster 1 - Cluster 2 | age | -0.27 | 0.13 | -2.01 | 0.11 |
| Cluster 1 - Cluster 3 | age | -0.49 | 0.13 | -3.74 | **<0.01** |
| Cluster 2 - Cluster 3 | age | -0.22 | 0.12 | -1.83 | 0.16 |
| Cluster 1 - Cluster 2 | c-T_chPain | -0.10 | 0.13 | -0.74 | 0.74 |
| Cluster 1 - Cluster 3 | c-T_chPain | -0.61 | 0.13 | -4.64 | **<0.01** |
| Cluster 2 - Cluster 3 | c-T_chPain | -0.51 | 0.12 | -4.18 | **<0.01** |
| Cluster 1 - Cluster 2 | COPC_head | 0.06 | 0.13 | 0.46 | 0.89 |
| Cluster 1 - Cluster 3 | COPC_head | -0.79 | 0.13 | -5.99 | **<0.01** |
| Cluster 2 - Cluster 3 | COPC_head | -0.85 | 0.12 | -6.96 | **<0.01** |
| Cluster 1 - Cluster 2 | H-CV-K_comb | 0.16 | 0.13 | 1.23 | 0.44 |
| Cluster 1 - Cluster 3 | H-CV-K_comb | -0.47 | 0.13 | -3.58 | **<0.01** |
| Cluster 2 - Cluster 3 | H-CV-K_comb | -0.64 | 0.12 | -5.20 | **<0.01** |
| Cluster 1 - Cluster 2 | low_BL | -0.63 | 0.13 | -4.75 | **<0.01** |
| Cluster 1 - Cluster 3 | low_BL | -0.66 | 0.13 | -5.00 | **<0.01** |
| Cluster 2 - Cluster 3 | low_BL | -0.02 | 0.12 | -0.20 | 0.98 |
| Cluster 1 - Cluster 2 | medicaid | -0.70 | 0.13 | -5.28 | **<0.01** |
| Cluster 1 - Cluster 3 | medicaid | -0.32 | 0.13 | -2.42 | **0.04** |
| Cluster 2 - Cluster 3 | medicaid | 0.39 | 0.12 | 3.15 | **<0.01** |
| Cluster 1 - Cluster 2 | pain_medication | -0.51 | 0.13 | -3.82 | **<0.01** |
| Cluster 1 - Cluster 3 | pain_medication | -0.86 | 0.13 | -6.56 | **<0.01** |
| Cluster 2 - Cluster 3 | pain_medication | -0.35 | 0.12 | -2.90 | **0.01** |
| Cluster 1 - Cluster 2 | tabaco | 0.25 | 0.13 | 1.89 | 0.14 |
| Cluster 1 - Cluster 3 | tabaco | -0.21 | 0.13 | -1.61 | 0.24 |
| Cluster 2 - Cluster 3 | tabaco | -0.47 | 0.12 | -3.80 | **<0.01** |
| Cluster 1 - Cluster 2 | unknown_race | -0.70 | 0.13 | -5.25 | **<0.01** |
| Cluster 1 - Cluster 3 | unknown_race | -0.35 | 0.13 | -2.67 | **0.02** |
| Cluster 2 - Cluster 3 | unknown_race | 0.35 | 0.12 | 2.84 | **0.01** |

| **Supplementary Table 4.** ANOVA table of the association of Phenotype with outcome (univariable analysis) for patients with 80% probability of membership or higher | | | | | |
| --- | --- | --- | --- | --- | --- |
|  | Df | Sum Sq | Mean Sq | F value | P value |
| **Delta PROMIS Physical (n = 218)** | | | | | |
| Phenotype | 2 | 136.4 | 68.21 | 1.247 | 0.28 |
| Residuals | 215 | 11752.1 | 54.66 |  |  |
| **Delta PROMIS Mental (n = 217)** | | | | | |
| Phenotype | 2 | 400.7 | 200.36 | 2.92 | 0.055 |
| Residuals | 214 | 14641.6 | 68.419 |  |  |
| **Delta VAS (n = 204)** | | | | | |
| Phenotype | 2 | 21.59 | 10.793 | 1.0618 | 0.34 |
| Residuals | 201 | 2043.24 | 10.165 |  |  |

**Supplementary Table 5:** ANOVA Table of parsimonious model for phenotyping factors predicting delta PROMIS Physical

| **Phenotyping factor** | **Df** | **Sum Sq** | **Mean Sq** | **F value** | **Adj. P value** |
| --- | --- | --- | --- | --- | --- |
| weight | 1 | 0.64 | 0.64 | 0.02 | 0.90 |
| age | 1 | 30.40 | 30.40 | 0.76 | 0.48 |
| male | 1 | 147.47 | 147.47 | 3.68 | 0.11 |
| COPC_head | 1 | 418.99 | 418.99 | 10.46 | **0.01** |
| c-T_chPain | 1 | 27.24 | 27.24 | 0.68 | 0.48 |
| unknown_race | 1 | 0.96 | 0.96 | 0.02 | 0.90 |
| leg_chPain | 1 | 269.52 | 269.52 | 6.73 | **0.04** |
| liver_disease | 1 | 203.30 | 203.30 | 5.08 | 0.07 |
| low_BL | 1 | 1297.06 | 1297.06 | 32.40 | **p<0.01** |
| tabaco | 1 | 63.41 | 63.41 | 1.58 | 0.28 |
| PT_visit | 1 | 13.55 | 13.55 | 0.34 | 0.62 |
| sciatica | 1 | 200.32 | 200.32 | 5.00 | 0.07 |
| COPD | 1 | 265.51 | 265.51 | 6.63 | **0.04** |
| asian | 1 | 289.60 | 289.60 | 7.23 | **0.04** |
| class | 2 | 160.46 | 80.23 | 2.00 | 0.21 |
| weight:class | 2 | 199.68 | 99.84 | 2.49 | 0.15 |
| unknown_race:class | 2 | 250.32 | 125.16 | 3.13 | 0.11 |
| leg_chPain:class | 2 | 229.50 | 114.75 | 2.87 | 0.11 |
| low_BL:class | 2 | 152.46 | 76.23 | 1.90 | 0.21 |
| COPD:class | 2 | 182.90 | 91.45 | 2.28 | 0.17 |
| asian:class | 2 | 648.43 | 324.22 | 8.10 | **p<0.01** |
| Residuals | *211* | *8448.04* | *40.04* |  |  |

**Supplementary Table 6:** ANOVA Table of parsimonious model for phenotyping factors predicting delta PROMIS Mental Health

| **Phenotyping factor** | **Df** | **Sum Sq** | **Mean Sq** | **F value** | **Adj. P value** |
| --- | --- | --- | --- | --- | --- |
| COPC_head | 1 | 702.02 | 702.02 | 14.25 | **p<0.01** |
| c-T_chPain | 1 | 0.22 | 0.22 | 0.00 | 0.99 |
| unknown_race | 1 | 5.94 | 5.94 | 0.12 | 0.81 |
| liver_disease | 1 | 329.28 | 329.28 | 6.69 | 0.07 |
| low_BL | 1 | 1505.67 | 1505.67 | 30.57 | **p<0.01** |
| LBP_chronicity | 1 | 155.13 | 155.13 | 3.15 | 0.14 |
| tabaco | 1 | 150.98 | 150.98 | 3.07 | 0.14 |
| sciatica | 1 | 186.04 | 186.04 | 3.78 | 0.14 |
| bipolar | 1 | 120.04 | 120.04 | 2.44 | 0.16 |
| COPD | 1 | 0.00 | 0.00 | 0.00 | 0.99 |
| other_dorsalgia | 1 | 15.12 | 15.12 | 0.31 | 0.69 |
| trunk_pain | 1 | 209.13 | 209.13 | 4.25 | 0.13 |
| asian | 1 | 169.58 | 169.58 | 3.44 | 0.14 |
| class | 2 | 337.27 | 168.63 | 3.42 | 0.13 |
| c-T_chPain:class | 2 | 192.14 | 96.07 | 1.95 | 0.18 |
| unknown_race:class | 2 | 230.51 | 115.25 | 2.34 | 0.14 |
| low_BL:class | 2 | 264.74 | 132.37 | 2.69 | 0.14 |
| COPD:class | 2 | 330.47 | 165.24 | 3.36 | 0.13 |
| other_dorsalgia:class | 2 | 243.40 | 121.70 | 2.47 | 0.14 |
| Residuals | 212 | 10440.96 | 49.25 |  |  |

**Supplementary Table 7:** ANOVA Table of parsimonious model for phenotyping factors predicting delta VAS

| **Phenotyping factor** | **Df** | **Sum Sq** | **Mean Sq** | **F value** | **Adj. P value** |
| --- | --- | --- | --- | --- | --- |
| weight | 1 | 0.60 | 0.60 | 0.08 | 0.78 |
| male | 1 | 22.42 | 22.42 | 2.90 | 0.14 |
| c-T_chPain | 1 | 1.94 | 1.94 | 0.25 | 0.65 |
| medicaid | 1 | 60.69 | 60.69 | 7.84 | **0.04** |
| H-CV-K_comb | 1 | 10.70 | 10.70 | 1.38 | 0.32 |
| mood_disorder | 1 | 22.15 | 22.15 | 2.86 | 0.14 |
| L-IVD_displ | 1 | 48.23 | 48.23 | 6.23 | **0.04** |
| tabaco | 1 | 3.70 | 3.70 | 0.48 | 0.54 |
| PT_visit | 1 | 25.01 | 25.01 | 3.23 | 0.14 |
| other_dorsalgia | 1 | 8.69 | 8.69 | 1.12 | 0.34 |
| L-IVD_deg | 1 | 84.87 | 84.87 | 10.96 | **0.02** |
| trunk_pain | 1 | 23.10 | 23.10 | 2.98 | 0.14 |
| asian | 1 | 48.67 | 48.67 | 6.29 | **0.04** |
| class | 2 | 55.52 | 27.76 | 3.59 | 0.07 |
| weight:class | 2 | 21.03 | 10.51 | 1.36 | 0.32 |
| male:class | 2 | 28.26 | 14.13 | 1.83 | 0.23 |
| c-T_chPain:class | 2 | 52.72 | 26.36 | 3.41 | 0.08 |
| tabaco:class | 2 | 56.61 | 28.30 | 3.66 | 0.07 |
| PT_visit:class | 2 | 70.98 | 35.49 | 4.58 | **0.04** |
| L-IVD_deg:class | 2 | 80.52 | 40.26 | 5.20 | **0.04** |
| Residuals | 194 | 1501.94 | 7.74 |  |  |

| **Supplementary Table 8. ANOVA Table of parsimonious model for intervention factors predicting delta PROMIS Physical (multivariable analysis)** | | | | | |
| --- | --- | --- | --- | --- | --- |
| Intervention factor | Df | Sum Sq | Mean Sq | F value | P value |
| ED_visit | 1 | 341.38 | 341.38 | 6.17 | **0.01** |
| *Residuals* | *238* | *13158.40* | *55.29* |  |  |

**Supplementary table 9:** ANOVA Table of parsimonious model for intervention factors predicting delta PROMIS Mental Health

| **Phenotyping factor** | **Df** | **Sum Sq** | **Mean Sq** | **F value** | **Adj. P value** |
| --- | --- | --- | --- | --- | --- |
| t_ED_visit | 1 | 293.41 | 293.41 | 4.64 | 0.10 |
| t_PT_visit | 1 | 0.71 | 0.71 | 0.01 | 0.92 |
| class | 2 | 363.34 | 181.67 | 2.87 | 0.10 |
| t_PT_visit:class | 2 | 332.77 | 166.39 | 2.63 | 0.10 |
| Residuals | 231 | 14598.40 | 63.20 |  |  |

**Supplementary table 10**: ANOVA Table of parsimonious model for intervention factors predicting delta VAS

| **Phenotyping factor** | **Df** | **Sum Sq** | **Mean Sq** | **F value** | **Adj. P value** |
| --- | --- | --- | --- | --- | --- |
| t_antidepressant | 1 | 58.43 | 58.43 | 6.31 | 0.09 |
| t_nerve_test | 1 | 25.87 | 25.87 | 2.80 | 0.11 |
| t_pain_medication | 1 | 31.89 | 31.89 | 3.45 | 0.11 |
| t_ED_visit | 1 | 26.94 | 26.94 | 2.91 | 0.11 |
| class | 2 | 40.48 | 20.24 | 2.19 | 0.11 |
| t_antidepressant:class | 2 | 45.67 | 22.83 | 2.47 | 0.11 |
| t_pain_medication:class | 2 | 46.87 | 23.44 | 2.53 | 0.11 |
| Residuals | 211 | 1952.21 | 9.25 |  |  |

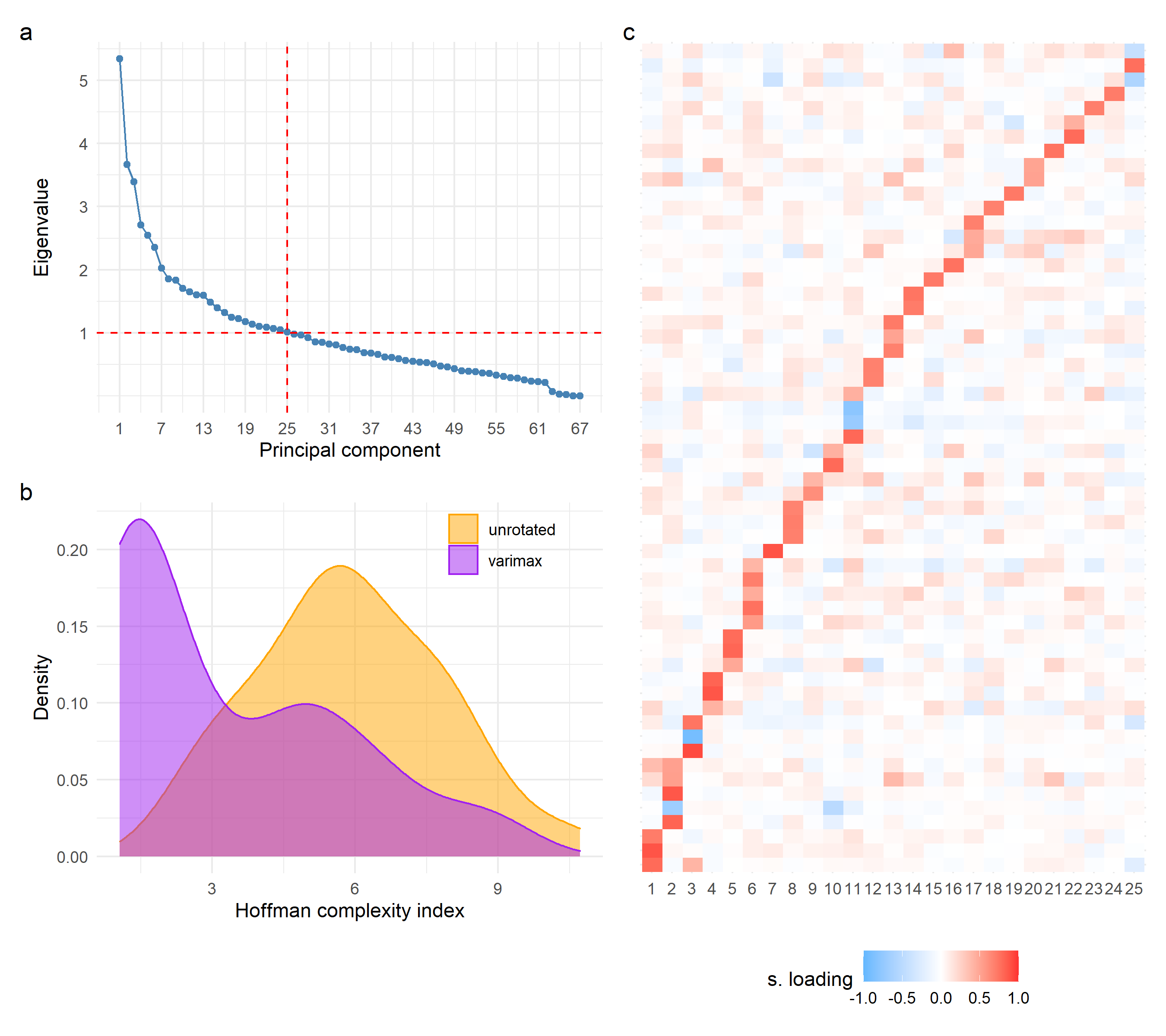

**Supplementary figure 1.** **Phenotyping factors extracted by NLPCA and varimax rotation.** The first 25 PCs had eigenvalues above 1 as shown in the scree plot (**a**). These were selected for further analysis following the Kaiser rule. Hoffman complexity index was used to measure loading complexity in the unrotated (initial solution) and the varimax NLPCA solutions (**b**). A heatmap of the standardized loadings after varimax rotation for the retained PCs (**c,** rows are variables and columns are PCs) shows how the rotated PCs are explained by a small subset of redundant variables. These rotated PCs are regarded as phenotyping factors.

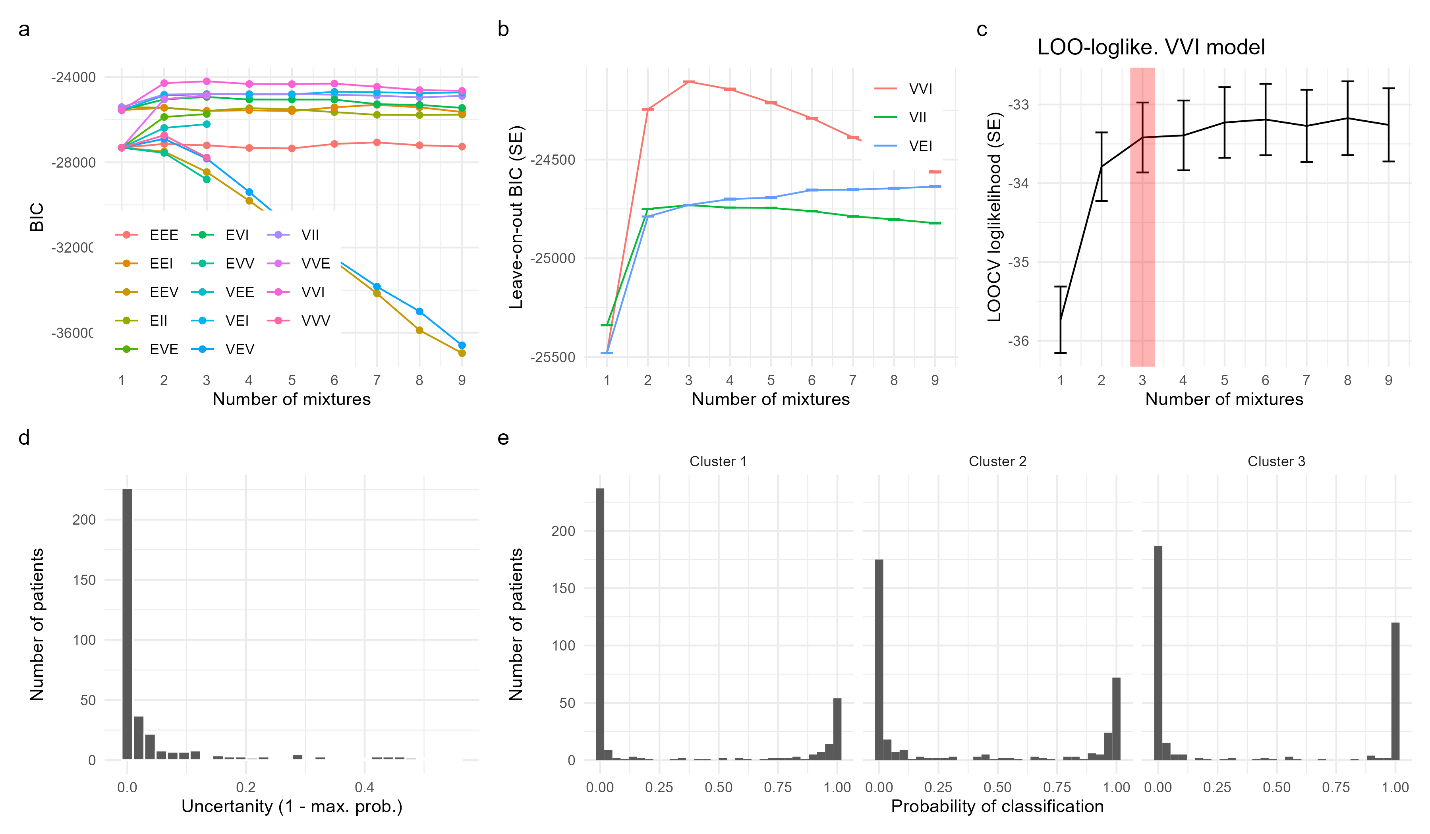

**Supplementary figure 2.** Selection of the best GMM covariance structure and number of mixtures (clusters). The different models with number of mixtures and covariance structure (see methods for definition) are compared based on BIC (**a**). The best three models (VVI, VII and VEI) where selected and a leave-on-out cross-validation (LOOCV) analysis was performed (**b**). This clearly show that a 3 clusters with VVI covariance structure fits the data best. We further examined the LOO loglikelihood of prediction accuracy (**c**) which shows that after 3 mixtures, adding more mixtures do not provide substantial gain in prediction of the LOO. The level of uncertainty for patient classification into clusters (**d**) and the probability of classification per cluster (**e**) show high confidence of cluster membership assignment (see Table 2 for summary).

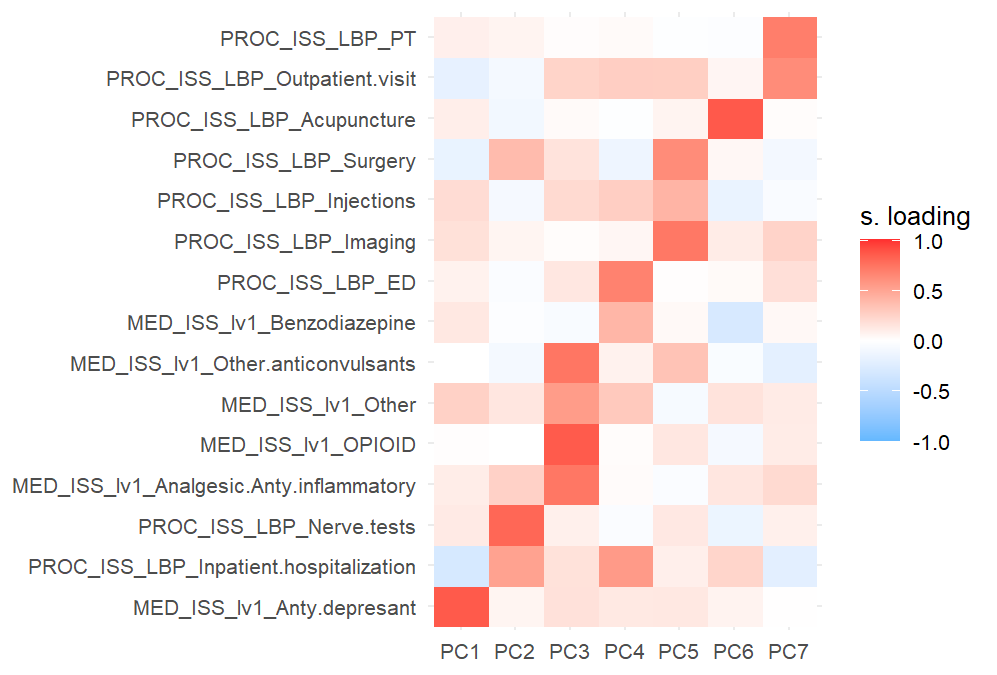

**Supplementary Figure 3.** Varimax rotated loadings of NLPCA from the “intervention” variables.

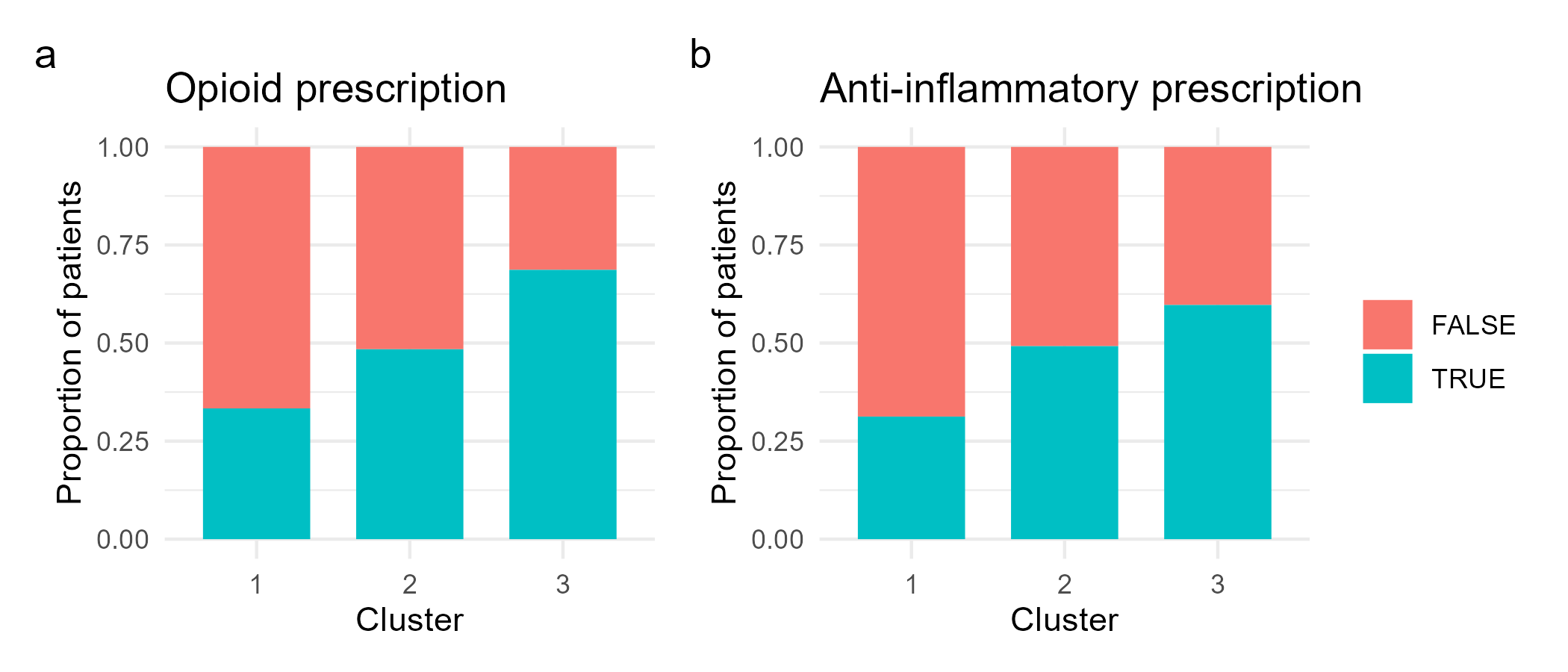

**Supplementary Figure 4.** Proportion of subjects prescived with opioids (**a**) and ani-inflammatory (**b**) drugs during treatment at ISS.

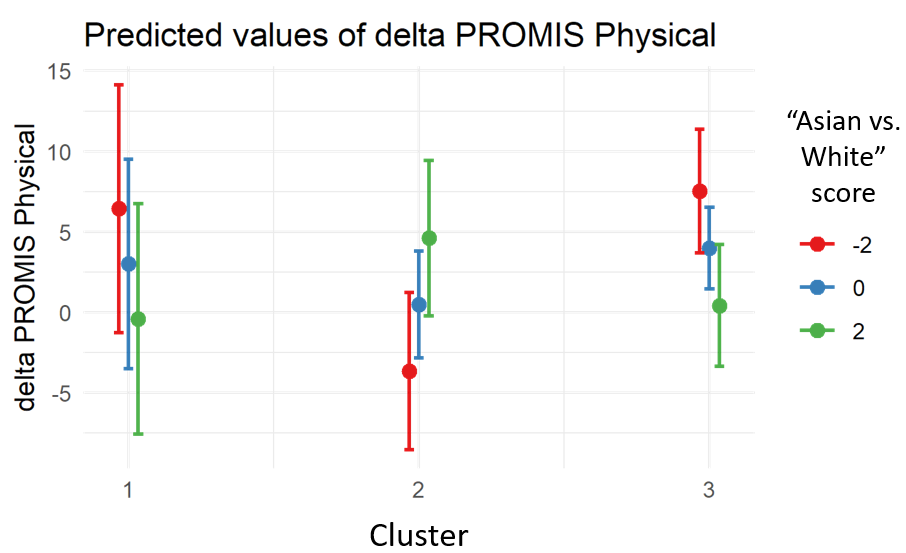

**Supplementary Figure 5.** Interaction plot of the “Asian vs. White” score and Cluster membership in the association of delta PROMIS Physical. The more positive the score is, the higher the probability that the patient is Asian, while the more negative the score is, the higher the probability that the patient is White.

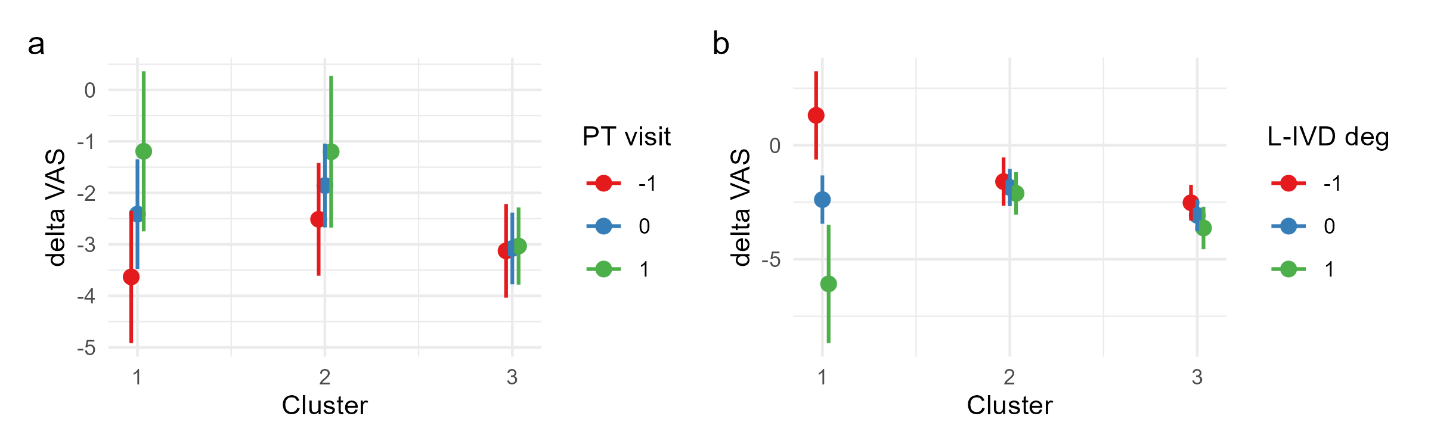

**Supplementary Figure 6.** Interaction plot of the “PT visits for LBP” score (a) and "Lumbar IVD degenerations" (b) with Cluster membership in the association of delta VAS.

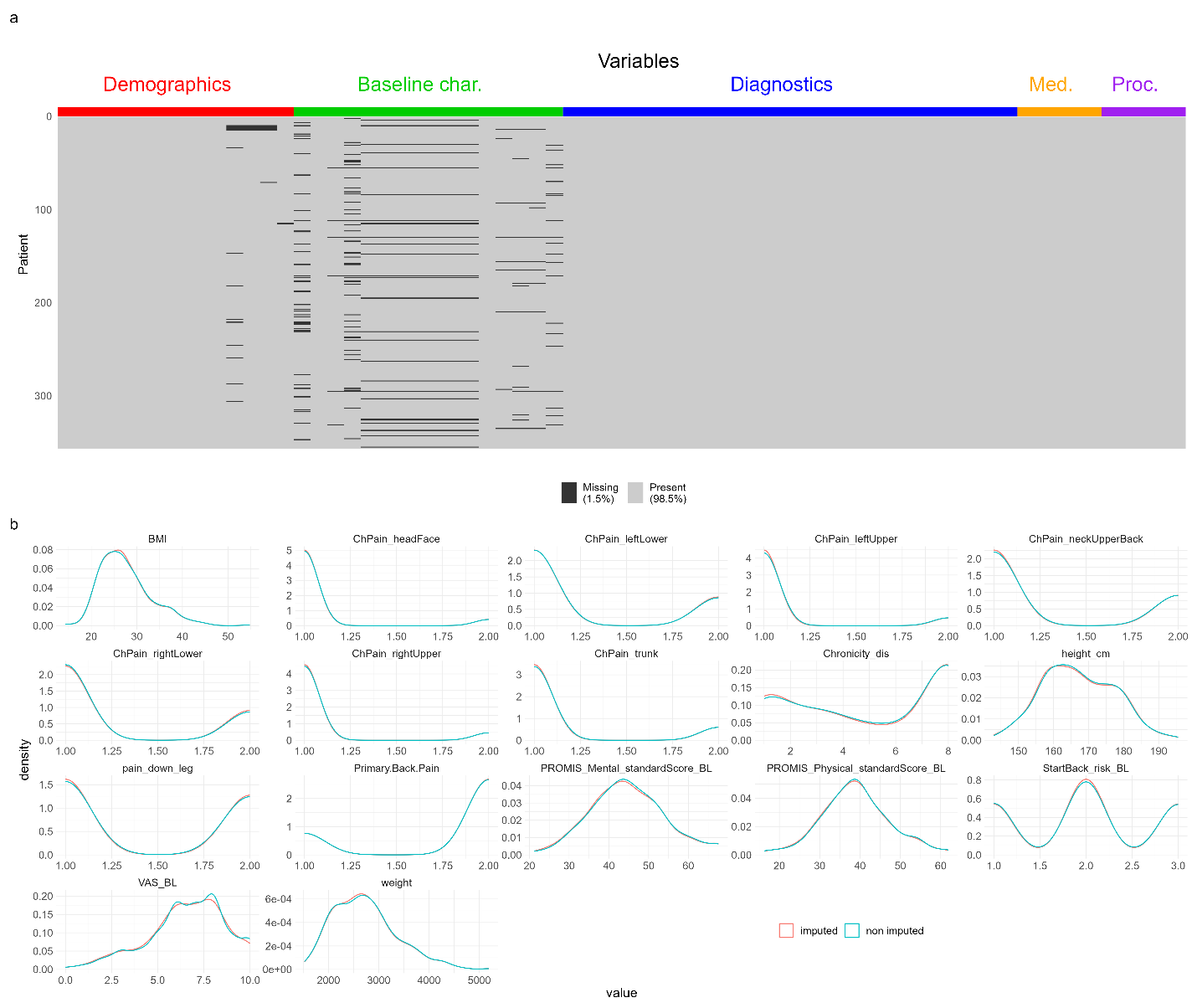

**Supplementary Figure 7.** Missing values analysis and imputation diagnostics. A shadow plot (**a**) shows the level of missing values mostly from the demographics and baseline characteristics variables. Multiple imputation was performed and the average distribution of the imputed vs. non imputed values used to determine any potential bias introduced buy the imputation process (**b**).
